# Development and validation of a novel circulating fibroblast activation protein - based predictive model to improve fibrosis risk stratification in metabolic liver disease population

**DOI:** 10.1101/2024.07.19.24310730

**Authors:** Ziqi V Wang, Badwi B Boumelhem, Torsten Pennell, William W Bachovchin, Jack Hung-Sen Lai, Sarah E Poplawski, Pieter Van Der Veken, Kate Brewer, Geraldine Ooi, Jacob George, Mohamed Eslam, Leon A Adams, Hui Emma Zhang, Geoffrey W McCaughan, Avik Majumdar, Mark D Gorrell

**Author notes:** Equal senior (Avik Majumdar, Mark D Gorrell).

## Abstract

**Objective:** Metabolic fatty liver disease drives chronic liver injury leading to fibrosis. This study aimed to establish a model utilising serum circulating fibroblast activation protein (cFAP) to diagnose advanced fibrosis in patients with fatty liver disease.

**Design:** Two retrospective cohorts recruited from tertiary hepatology clinics were studied as training (n=160) and external validation cohorts (n=342), with prevalence of histologic advanced fibrosis (F3-F4) of 20% and 11%, respectively. A marker of activated mesenchymal fibrogenic cells, cFAP, was measured using our single-step enzyme assay. A predictive model, FAP Index, containing age, type 2 diabetes, alanine transaminase and ordinal cFAP was developed using logistic regression. Diagnostic accuracy of FAP Index was assessed on a single and then sequential basis.

**Results:** FAP Index AUROC was 0.875 (95% CI 0.813-0.938) in the training cohort and 0.841 (95% CI 0.776-0.906) in the validation cohort. Low cut-off −1.68 (Sensitivity 80.0%, negative predictive value 95.5%) and high cut-off +0.953 values (Specificity 97.7%, positive predictive value 88.9%) excluded and diagnosed advanced fibrosis, respectively. In the validation cohort, FAP Index then FIB-4 reduced indeterminate results by one-third compared to FIB-4 alone. Whereas FAP Index followed by NFS (NAFLD Fibrosis Score) resulted in a reduction of indeterminate results by 70% compared to NFS alone.

**Conclusion:** FAP Index is a novel, rapid, robust, inexpensive diagnostic tool for advanced fibrosis in metabolic fatty liver disease. Applying FAP Index followed by FIB-4 or NFS facilitates accurate risk-stratification of patients by greatly reducing the frequency of indeterminate results compared to FIB-4 or NFS alone, without compromising negative predictive value.

**What is already known on this topic:** Fatty liver disease affects one quarter of the global population. Current screening algorithms to triage those at high risk of advanced fibrosis use a dual cut-off approach that results in a proportion of patients that cannot be classified (indeterminate result) and hence need further and more costly testing.

**What this study adds:** We have developed the FAP Index, which is a model using a simple circulating fibroblast activation protein enzyme assay and routinely available clinical variables. Using FAP Index as a first-line test followed by the current recommended screening tests (FIB-4 and NFS [NAFLD Fibrosis Score]) can reduce indeterminate results by up to 70% compared to the current first-line standard of care tests alone, without compromising diagnostic accuracy.

**How this study might affect research, practice or policy:** With recently approved pharmacotherapy for fatty liver disease, improved tools for triaging people with metabolic fatty liver disease has increasing urgency. Use of FAP Index could have a dramatic effect on screening for advanced fibrosis by reducing fruitless referrals to tertiary care and/or further testing. Furthermore, our single-step enzymatic cFAP assay can be adapted to point of care or reflex testing settings, allowing for low-cost and high throughput FAP Index screening.

## 1 Introduction

Fatty liver disease is becoming an increasingly common metabolic disorder and currently affects one quarter of the global population^1, 2^. The pathogenesis of fatty liver disease encompasses multi-factorial causes such as abnormal lipid metabolism, glucose dysregulation, gut microbiome alterations and genetic variants^3^. These abnormal physiological changes promote chronic inflammation that can lead to hepatic fibrosis, which is the most potent driver towards end-stage liver disease and hepatocellular carcinoma. Moreover, obesity, type 2 diabetes mellitus (T2DM) and insulin resistance drive this disease^4, 5^. Early-stage metabolic fatty liver disease and hepatic fibrosis is usually asymptomatic^6, 7^. However, the risk of liver-related morbidity and mortality increases dramatically in parallel with the progression into advanced hepatic fibrosis and cirrhosis^8, 9^. As the presence of advanced fibrosis is the major determinant of liver-related complications, accurately identifying those at risk in the context of increasing metabolic associated fatty liver disease prevalence has become an urgent clinical need for hepatologists, diabetologists and primary care physicians.

Histological assessment with liver biopsy is becoming less frequently used due to its invasive and inherent sampling bias. New 3D imaging techniques highlight the latter limitation by starkly revealing the uneven localisation of fibrosis^10^. Elastography techniques are not widely accessible in the community. Therefore, non-invasive blood tests have come to the forefront for the diagnosis of advanced liver fibrosis in patients with metabolic disorders. The two most commonly used models are the NAFLD fibrosis score (NFS)^11^ and the Fibrosis index-4 (FIB-4)^12^. However, despite being recommended for advanced fibrosis screening, these tests require further optimisation^13, 14^. Recently, the European Association for the Study of the Liver (EASL) has proposed a three-step strategy including non-invasive tests (NITs) to screen for advanced fibrosis^15^. This algorithm has been further validated for discriminating advanced fibrosis, where it displays superior accuracy to a single test algorithm^8^. Despite the limitations of these NITs, the application of low-cost and simple sequential NITs appears more appropriate to screening in the community setting than higher cost alternatives.

Fibroblast activation protein alpha (FAP) is a dimeric type 2 trans-membrane glycoprotein with proteolytic activity that degrades fibroblast growth factor 21 (FGF21)^16, 17^, neuropeptide Y^18^ and denatured collagens^17, 19, 20^, and activates α2-antiplasmin^19^. FAP is near-absent from healthy tissues but highly expressed by activated stellate cells and myofibroblasts in cirrhosis^20–22^. Moreover, FAP is heavily glycosylated (∼30% by mass), which may confer resistance to degradation that explains the persistence of soluble FAP in circulation (cFAP)^23^. We previously reported our specific enzyme assay of FAP^24, 25^, and have shown that increased cFAP activity associates with advanced fibrosis (F3-F4)^25^. Additionally, cFAP activity decreases following liver transplantation^26^. Thus, cFAP activity is a potential serum biomarker for liver fibrosis that derives from the cell types that produce and regulate most of the liver extracellular matrix (ECM)^17, 19^. Moreover, FAP colocalizes with ECM proteins in liver fibrosis^22, 27, 28^. Thus, it is important to explore the utility of measuring FAP when assessing fibrosis. FAP can be measured by multiple approaches including specific substrate catalytic assay, antibody capture catalytic assay or ELISA^24, 29–31^. Our robust, single-step in-house FAP assay that uses a FAP-specific fluorescent substrate^24^ strongly correlates with the commercial ELISA. ^24, 26, 29–31^

In this study, the primary aim was to develop a NIT-based fibrosis algorithm that incorporates our specific, quantitative cFAP enzyme activity assay in patients with metabolic associated fatty liver disease to identify advanced fibrosis, and then to validate the performance of this algorithm upon a cohort that mimics a community population. We also examined whether the two most prominent FAP-specific substrates^24, 29^ generate comparable quantitative data on FAP abundance in serum samples.

## 2 Method

### 2.1 Study cohorts

Two retrospective study cohorts were obtained from their originating hospitals. The training cohort (n=160) consisted of 65 patients who underwent liver biopsy for staging of fatty liver disease and 95 patients who had liver biopsy at the time of bariatric surgery at Sir Charles Gairdner Hospital, Perth, Australia and has been described in part previously^32^. The validation cohort (n=332) was pooled from two previously described study populations; 182 patients who had liver biopsy at the time of bariatric surgery at The Alfred Hospital, Melbourne, Australia^33^ and 150 patients who underwent liver biopsy for fibrosis staging of metabolic associated fatty liver disease (MAFLD) at Westmead Hospital, Sydney, Australia^34^. These cohorts were pooled in order to replicate the prevalence of advanced fibrosis in an at-risk screening population of MAFLD, which is estimated to be 10 to 15% in Australia^35, 36^. All included patients met MAFLD criteria^37^ and had available stored serum (−80°C) to perform cFAP assay. ^37^ The exclusion criteria were (1) alcohol related aetiology (average of >30 grams daily for men and >20 grams daily for women), (2) chronic viral hepatitis and (3) non-metabolic forms of liver disease. Ethics approvals were HREC_X18-0241 in Sydney Local Health District, 2019/ETH02319 in Westmead Hospital, RGS 01287 in Sir Charles Gairdner Hospital, and 195/15 in The Alfred Hospital.

### 2.2 Clinical, demographic and histologic data

Demographic data and routine clinical laboratory values (haematology and biochemistry) were obtained within four weeks of the liver biopsy and measured in standardized units used in Australia. Histological fibrosis assessments were performed for all included patients by the pathology service in each hospital where liver biopsy occurred, as previously described.^32–34^ Liver fibrosis grade was evaluated according to the Kleiner pathology scoring system.^38^ The outcome variable was advanced liver fibrosis, defined as fibrosis staging >2 (F3-F4). Data were de-identified.

### 2.3 The cFAP assay

The cFAP assay was performed as previously described^24^. Briefly, 5 µL of a 1:5 dilution of either human or mouse serum was pipetted into replicate wells of a 96-well plate and topped up to 70 µL with tris-acetate (10 mM)-EDTA (1 mM) (pH 7.4) (TE buffer). Serial dilutions of 7-amino-4-methylcoumarin (0-600 pmol) were pipetted in replicate wells to a final volume of 100 µL per well. The plate was read on a BMG PolarStar plate reader (BMG Labtech, Ortenberg, Germany) set up to read at excitation 355 nm & emission 450 nm every 2.5 mins for 1 hr at 37°C.

### 2.4 Statistical analysis

Data are presented as median with interquartile range (IQR) due to non-parametric distribution and means with standard deviation otherwise. Statistical tests were performed with exclusion of missing data for variables of interest. Mann-Whitney U, one-way ANOVA where appropriate. Linear correlations were assessed with Pearson’s correlation coefficient.

Univariable logistic regression was performed when the dependent variable was binary vs a continuous independent variable. ^39^ Chi-square test was performed when both dependent and independent variables were non-linear. The distribution of cFAP activity as a continuous variable revealed extreme outliers in the training cohort (figure S1). Therefore, cFAP activity was binned into 3 groups (ordinals): 0=low, 1=middle and 2=high using cut-offs at 730 and 1580 pmol AMC/min/mL. The low cut-off was as established by our group previously^25^, and the high cut-off was selected based on optimising specificity for advanced fibrosis. Both continuous and binned cFAP activity were tested for strength of association with advanced fibrosis by binary logistic regression.

To optimise diagnostic accuracy, a multivariable logistic regression model for advanced fibrosis incorporating cFAP and clinical variables was developed following published methods^39^. Variables were selected for multivariable analysis if p < 0.1 on univariable analysis. Backward elimination was then used to arrive at the final model. Interaction between predictors was not accounted in this study. The Hosmer-Lemeshow test was used to assess the goodness of fit of the final model, termed FAP Index. Area under the receiver operating characteristic curves (AUROC) were calculated for evaluating diagnostic accuracy of FAP Index in training and validation cohorts. An AUROC greater 0.7 was considered good while a score greater than 0.9 is outstanding^40^. Dual cut-off points were chosen from the ROC curve for FAP Index to optimize sensitivity and specificity in classifying patients into low- and high-risk of advanced fibrosis, respectively. Positive predictive value (PPV), negative predicted value (NPV)^41^, sensitivity, specificity and accuracy ^41^ were calculated without the indeterminate population, since that population remained ‘unclassified’. FAP index, was compared with FIB-4 ^12^ and NFS ^11^, both as standalone NITs and as part of sequential NIT combinations. Complete case analysis was applied upon model validation in both training and validation cohorts. FIB-4 and NFS scores were calculated in only 87 patients in the training cohort due to missing data (designated as the training sub-cohort), however, complete data were available for the validation cohort. Delong test was used to compare AUROCs. An alpha value of 0.05 was the threshold for statistical significance. Statistical analyses were performed in R studio (version 2023.03.0+386) and GraphPad Prism (GraphPad, v9.4.1).

## 3 Results

### 3.1 Clinical characteristics of study cohorts

Characteristics of study cohorts are presented in table 1 and supplemental table 1. There were no significant differences between the training cohort and the validation cohort except for age, the prevalence of fibrosis stages and cFAP level activities. The training cohort included more advanced fibrosis patients (20.3%) than the validation cohort (11.4%) (P = 0.01). Of note, there was no difference in the prevalence of type 2 diabetes (T2DM), which was 36% in the training and 30% in the validation cohort (P = 0.21). The median body mass index (BMI) also did not differ for the training and the validation cohorts at 38.19 (12.05) and 38.6 (14.2) kg/m^2^ respectively (P = 0.89). HOMA-IR is significantly greater in the validation cohort (2.49 (3.18)) compared to the training cohort (0.4 (0.76)) (P < 0.0001). The cFAP activity was greater in the validation cohort (1290 (555.9) pmol AMC/min/L) than the training cohort (995.74 (579.3) pmol AMC/min/L) (P < 0.0001). However, there were no significant differences observed for liver enzymes. Circulating FAP was found to associate with HOMA-IR, insulin and all three liver transaminases in both cohorts (P < 0.05; supplemental table 2), but cFAP did not associate with T2DM.

**Table 1.**
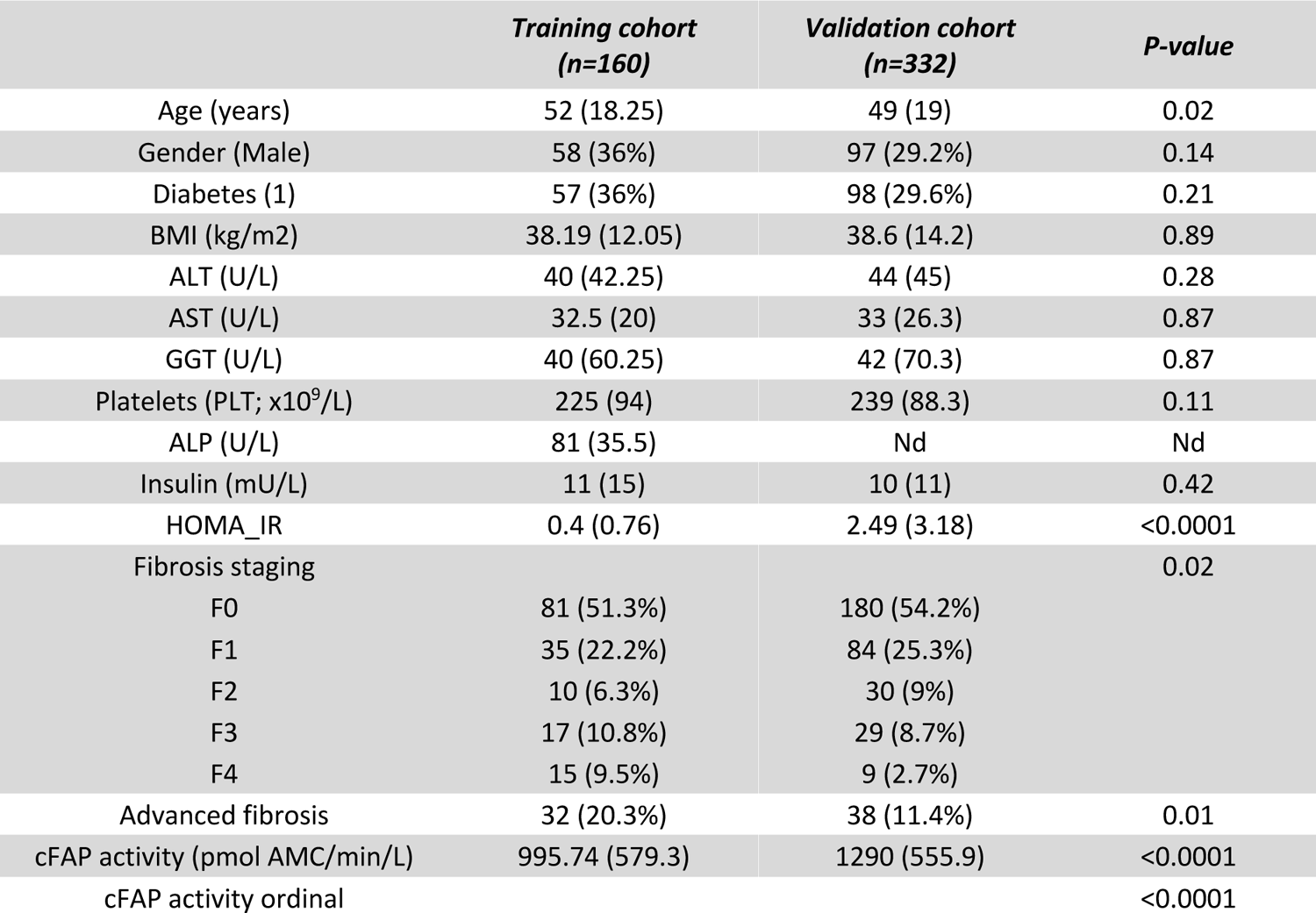
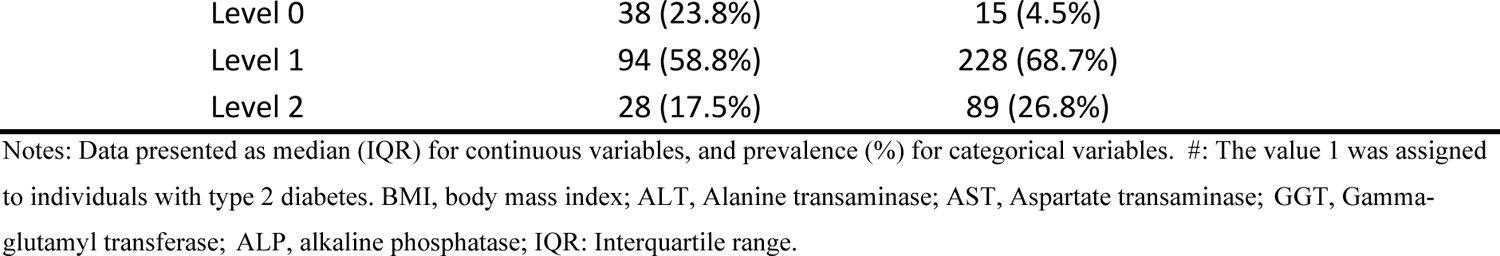
Baseline characteristics of training and validation cohort. Statistically significant differences were obtained by Mann-Whitney U test (P-value).

|  | Training cohort<br>(n=160) | Validation cohort<br>(n=332) | P-value |
| --- | --- | --- | --- |
| Age (years) | 52 (18.25) | 49 (19) | 0.02 |
| Gender (Male) | 58 (36%) | 97 (29.2%) | 0.14 |
| Diabetes (1) | 57 (36%) | 98 (29.6%) | 0.21 |
| BMI (kg/m<sup>2</sup>) | 38.19 (12.05) | 38.6 (14.2) | 0.89 |
| ALT (U/L) | 40 (42.25) | 44 (45) | 0.28 |
| AST (U/L) | 32.5 (20) | 33 (26.3) | 0.87 |
| GGT (U/L) | 40 (60.25) | 42 (70.3) | 0.87 |
| Platelets (PLT; x10<sup>9</sup>/L) | 225 (94) | 239 (88.3) | 0.11 |
| ALP (U/L) | 81 (35.5) | Nd | Nd |
| Insulin (mU/L) | 11 (15) | 10 (11) | 0.42 |
| HOMA_IR | 0.4 (0.76) | 2.49 (3.18) | <0.0001 |
| Fibrosis staging |  |  | 0.02 |
| F0 | 81 (51.3%) | 180 (54.2%) |  |
| F1 | 35 (22.2%) | 84 (25.3%) |  |
| F2 | 10 (6.3%) | 30 (9%) |  |
| F3 | 17 (10.8%) | 29 (8.7%) |  |
| F4 | 15 (9.5%) | 9 (2.7%) |  |
| Advanced fibrosis | 32 (20.3%) | 38 (11.4%) | 0.01 |
| cFAP activity (pmol AMC/min/L) | 995.74 (579.3) | 1290 (555.9) | <0.0001 |
| cFAP activity ordinal |  |  | <0.0001 |

|  |  |  |
| --- | --- | --- |
| Level 0 | 38 (23.8%) | 15 (4.5%) |
| Level 1 | 94 (58.8%) | 228 (68.7%) |
| Level 2 | 28 (17.5%) | 89 (26.8%) |

Notes: Data presented as median (IQR) for continuous variables, and prevalence (%) for categorical variables. #: The value 1 was assigned to individuals with type 2 diabetes. BMI, body mass index; ALT, Alanine transaminase; AST, Aspartate transaminase; GGT, Gamma-glutamyl transferase; ALP, alkaline phosphatase; IQR: Interquartile range.

### 3.2 FAP Enzyme assay specificity and reproducibility

The specificity of our assay for FAP ^24^ has been questioned ^29^ and so was revisited and reaffirmed (supplemental material S2.1; supplemental figure 2). Robust assay reproducibility following freeze/thaw cycles and prolonged ultracold storage were established (supplemental Material S2.2; supplemental figure 3), consistent with our previous data^24, 26^.

### 3.3 Application of cFAP for hepatic fibrosis detection

The cFAP activity was significantly associated with fibrosis (supplemental table 2). In particular, cFAP of patients with F3-F4 was significantly greater (P<0.0001) than patients without fibrosis (F0) in the training cohort (figure 1A-B), suggesting an ability to discriminate between absence of fibrosis and presence of advanced fibrosis. Significant differences in cFAP were also observed in inter-stage comparisons with moderate liver fibrosis in the validation cohort.

**Figure 1.**
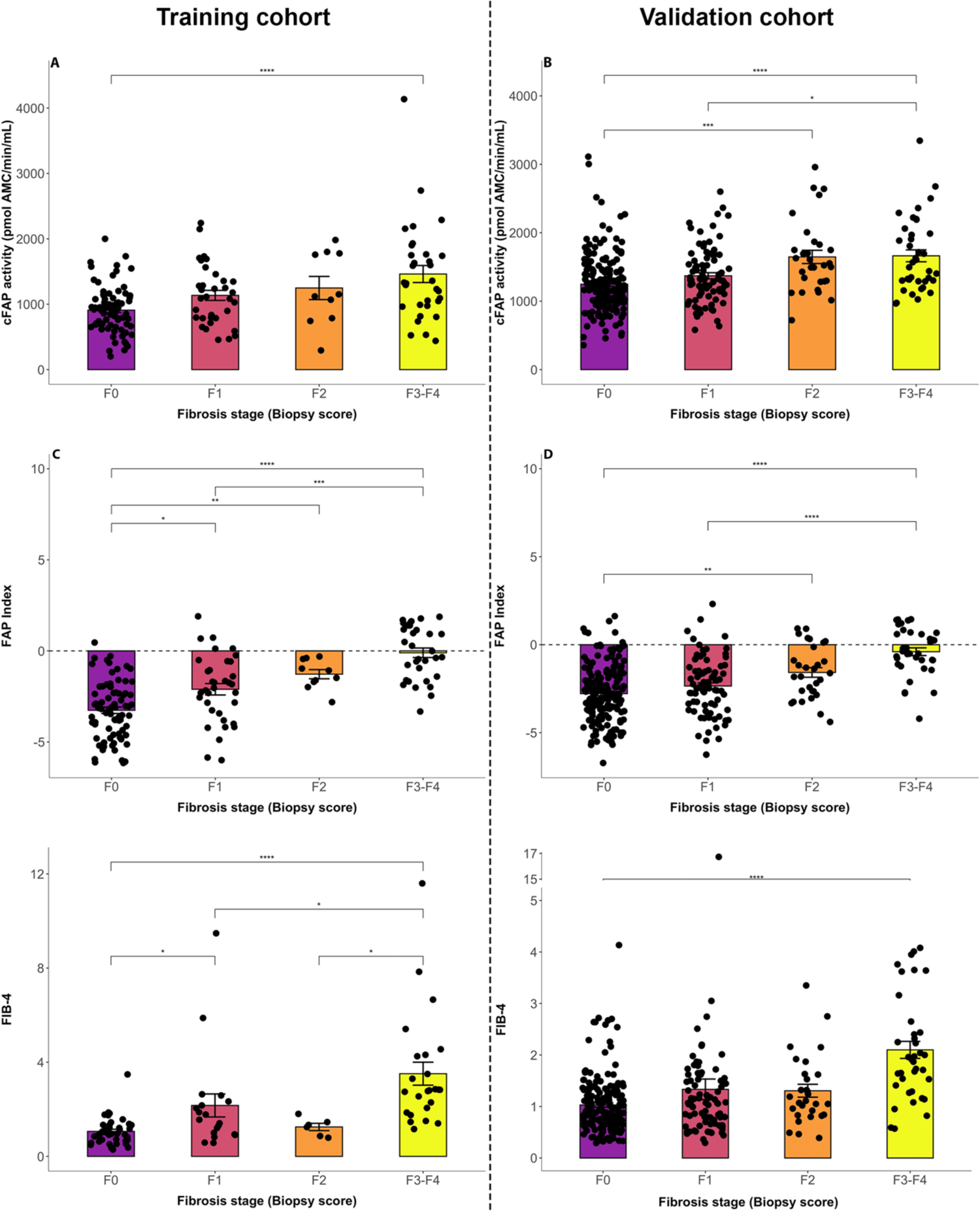
Associations of cFAP activity, FAP Index and FIB-4 with fibrosis staging. The cFAP activity (U/L) (A, B), FAP Index (C, D) and FIB-4 (E, F) segregated according to fibrosis stage are displayed for training cohort (n=160) (A, C, E) and validation cohort (n = 332) (B, D, F). Dot plots with mean ± SEM. Differences between groups were determined using a one-way ANOVA with Tukey’s post-hoc test. Significant differences are indicated with asterisks to indicate degree of difference: * p-value<0.05, ** p-value<0.01, *** p-value<0.001, **** p-value<0.0001.

### 3.4 Model development for the detection of advanced fibrosis

Multivariable logistic regression revealed age, T2DM, ALT and cFAP (table 2) as predictors of advanced fibrosis. We therefore derived FAP Index using the following formula:

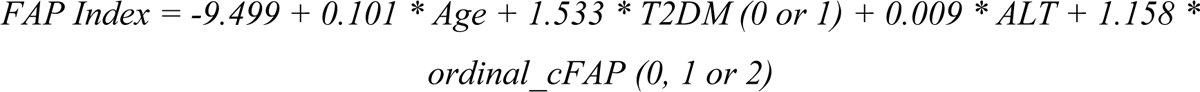

**Table 2.**
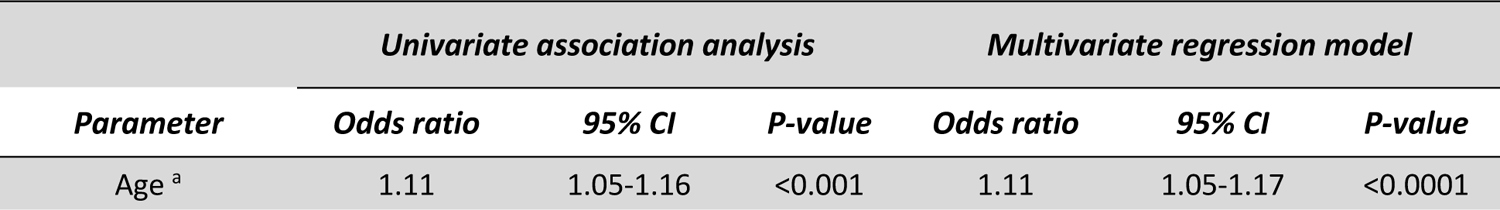
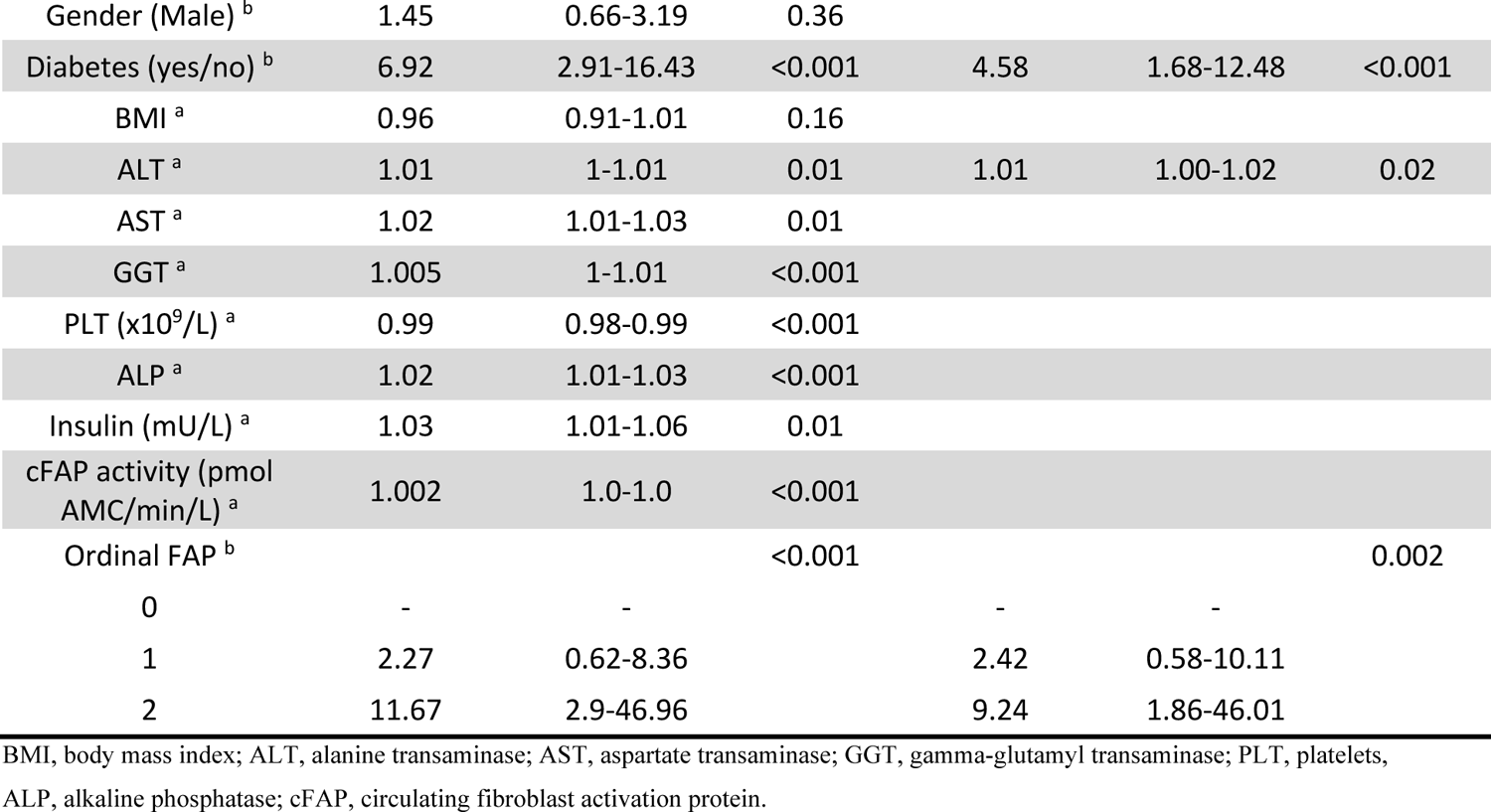
Associations between clinical parameters and advanced fibrosis in training cohort. ^a^ Univariate logistic regression; ^b^ Chi-square test.

| Parameter | Univariate association analysis |  |  | Multivariate regression model |  |  |
| --- | --- | --- | --- | --- | --- | --- |
|  | Odds ratio | 95% CI | P-value | Odds ratio | 95% CI | P-value |
| Age<sup>a</sup> | 1.11 | 1.05-1.16 | <0.001 | 1.11 | 1.05-1.17 | <0.0001 |
| Gender (Male) <sup>b</sup> | 1.45 | 0.66-3.19 | 0.36 |  |  |  |
| Diabetes (yes/no) <sup>b</sup> | 6.92 | 2.91-16.43 | <0.001 | 4.58 | 1.68-12.48 | <0.001 |
| BMI <sup>a</sup> | 0.96 | 0.91-1.01 | 0.16 |  |  |  |
| ALT <sup>a</sup> | 1.01 | 1-1.01 | 0.01 | 1.01 | 1.00-1.02 | 0.02 |
| AST <sup>a</sup> | 1.02 | 1.01-1.03 | 0.01 |  |  |  |
| GGT <sup>a</sup> | 1.005 | 1-1.01 | <0.001 |  |  |  |
| PLT (x10<sup>9</sup>/L) <sup>a</sup> | 0.99 | 0.98-0.99 | <0.001 |  |  |  |
| ALP <sup>a</sup> | 1.02 | 1.01-1.03 | <0.001 |  |  |  |
| Insulin (mU/L) <sup>a</sup> | 1.03 | 1.01-1.06 | 0.01 |  |  |  |
| cFAP activity (pmol<br>AMC/min/L) <sup>a</sup> | 1.002 | 1.0-1.0 | <0.001 |  |  |  |
| Ordinal FAP <sup>b</sup> |  |  | <0.001 |  |  | 0.002 |
| 0 | - | - |  | - | - |  |
| 1 | 2.27 | 0.62-8.36 |  | 2.42 | 0.58-10.11 |  |
| 2 | 11.67 | 2.9-46.96 |  | 9.24 | 1.86-46.01 |  |

BMI, body mass index; ALT, alanine transaminase; AST, aspartate transaminase; GGT, gamma-glutamyl transaminase; PLT, platelets, ALP, alkaline phosphatase; cFAP, circulating fibroblast activation protein.

The Hosmer-Lemeshow test χ^2^ was 3.066 (P = 0.93), indicating that FAP Index was well fitted. In the training cohort, the AUROC for FAP Index in predicting advanced fibrosis was 0.875 (95% CI: 0.813 – 0.938; figure 2A). Based on the ROC, a dual cut-off strategy was applied to optimise FAP Index for ruling in and out advanced fibrosis. The low cut-off was chosen at −1.681 (sensitivity = 0.84, specificity = 0.75), and the high cut-off at 0.953 (sensitivity = 0.34, specificity = 0.99). Patients with a score lying in between these low and high cut-off scores were designated indeterminate.

**Figure 2.**
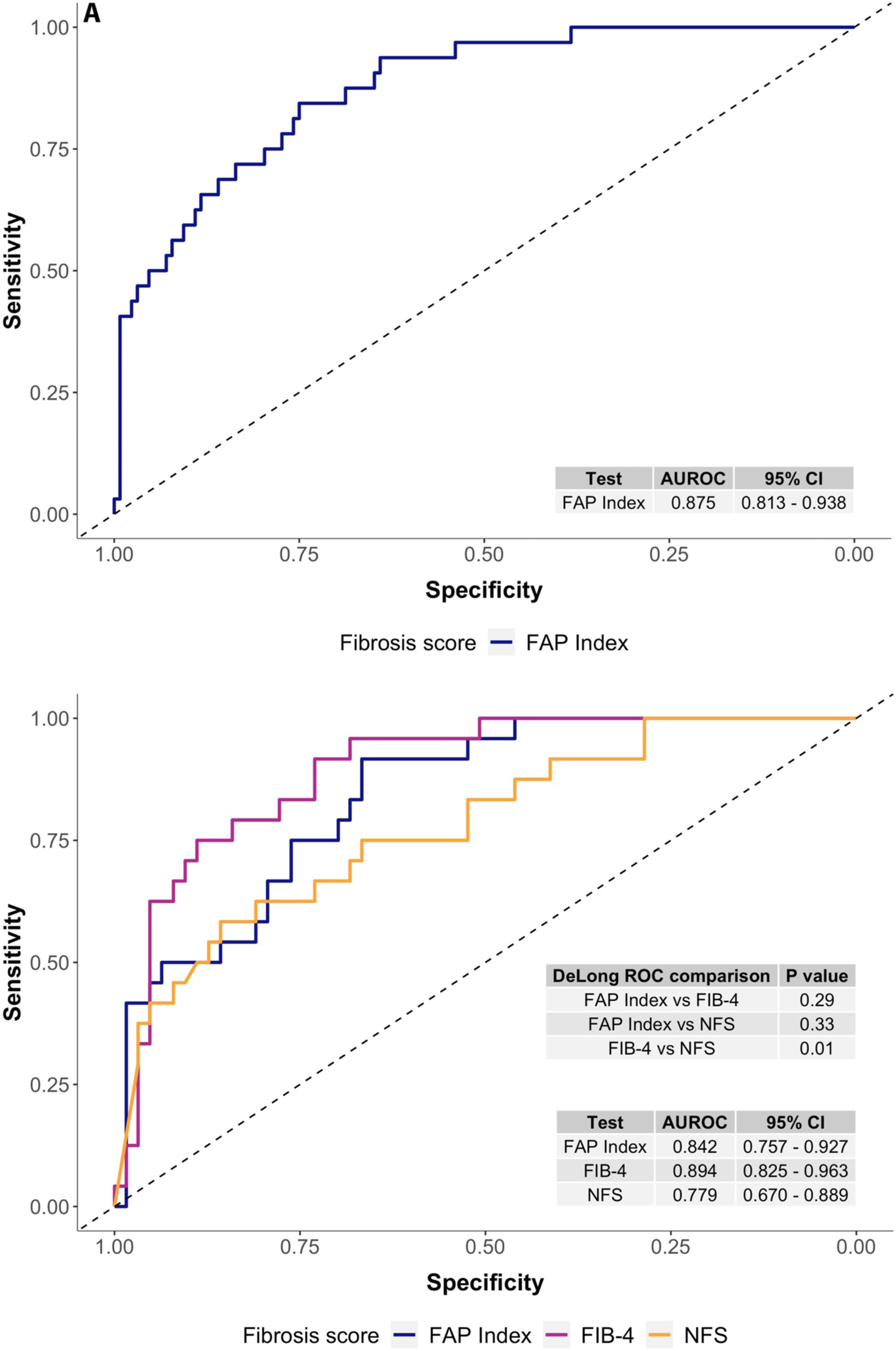
(A) The receiver operating characteristics curve for the FAP Index in the training cohort (AUROC = 0.875 with 95% CI: 0.757 – 0.927). (B) The AUROC for the FAP Index is comparable with FIB-4 and NFS. Receiver operating characteristics curve of test scores in the training cohort subset including FAP Index (AUROC = 0.842 with 95% CI: 0.757-0.927), FIB-4 (AUROC = 0.894 with 95% CI: 0.825-0.963) and NFS (AUROC = 0.779 with 95% CI: 0.67-0.889). Statistical significance between ROC curves was tested by DeLong test.

The association of FAP Index with fibrosis severity (figure 1C-D) displayed better differentiation across fibrosis stages than with cFAP alone and comparable with FIB-4 (figure 1E-F).

### 3.5 Comparison of FAP Index to existing NITs

Only patients with complete data for FAP Index, NFS and FIB-4 were included, which was designated as the training sub-cohort (n=87). The training sub-cohort was compared with its original cohort (supplemental table 2) and was older (57 vs 52, P=0.03), had lower BMI (34 vs 38, P = 0.02) and had higher insulin levels (15 vs 11, P=0.02) and cholestatic liver enzymes. Notably, there was no difference in the prevalence of advanced fibrosis. The FAP Index accuracy was numerically superior to NFS and similar to FIB-4 (AUROC 0.842, 0.779 and 0.894, respectively), but without a statistically significant difference between FAP Index and FIB-4 or NFS (figure 2B). The NPVs of FAP Index, NFS and FIB-4 were 95%, 92.6% and 95.9%, respectively, with corresponding specificities of 97.7%, 80.6% and 93.8%. The sensitivities of FAP Index, NFS and FIB-4 were between 80 and 85%. Use of FAP Index and FIB-4 as standalone tests resulted in 39.1% and 32.2% of patients classified as indeterminate, respectively, whereas NFS generated 49.4% indeterminate (figure 3A-C, table 3). FAP Index, when considered, as a continuous variable, correlated with both FIB-4 and NFS (P < 0.001; supplemental Material S2.3; supplemental figure 4)

**Figure 3.**
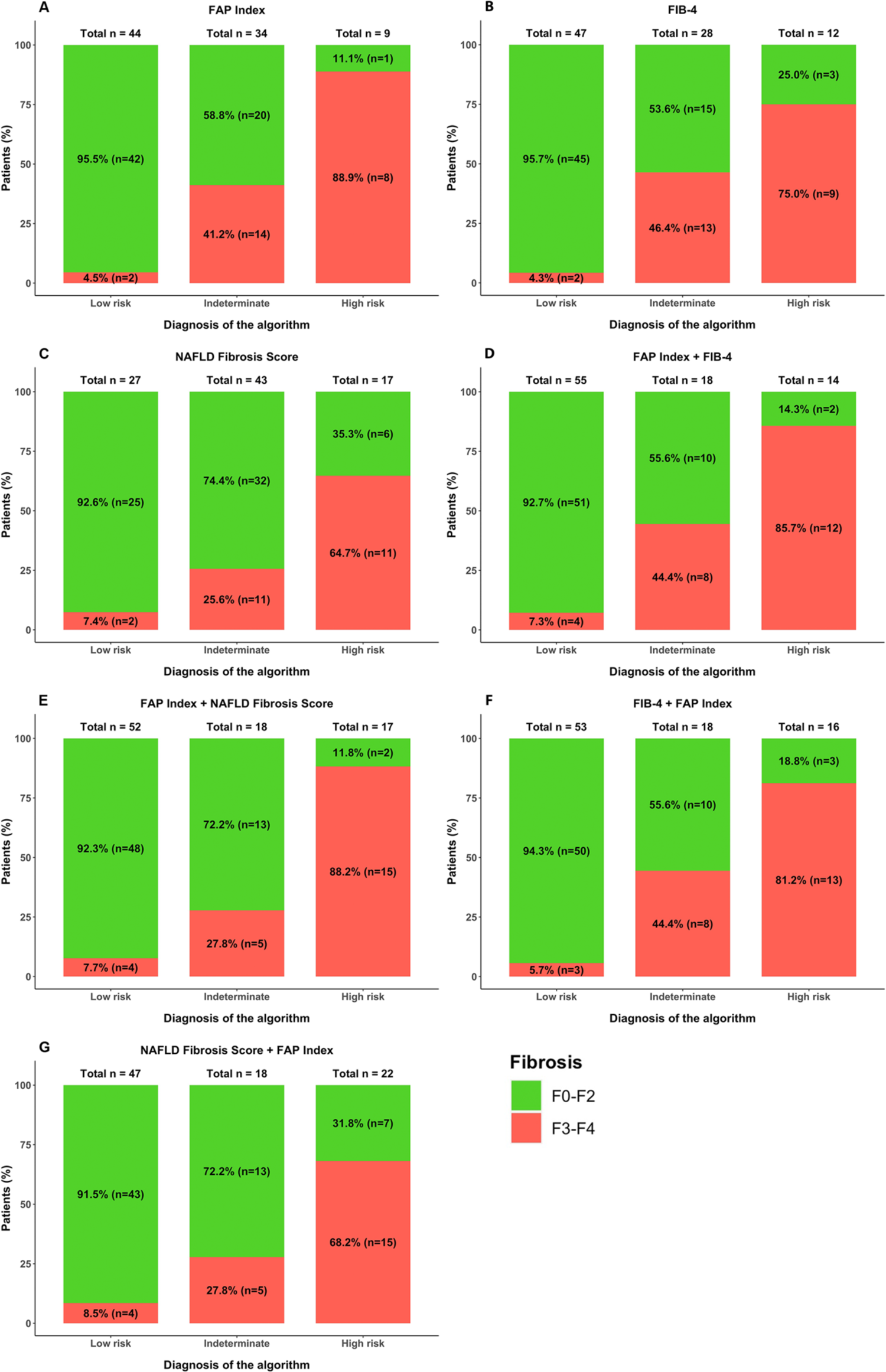
Classification analyses in training cohort. Fibrosis stages classification as single test or sequential tests, as a function of subgroups defined by: **A.** FAP Index classification. **B.** FIB-4. **C.** NAFLD Fibrosis Score. **D.** FAP Index followed by FIB-4. **E.** FAP Index followed by NAFLD Fibrosis Score. **F.** Fibrosis stages as a function of subgroups defined by FIB-4 followed by FAP Index. **G.** NAFLD Fibrosis Score followed by FAP Index.

**Table 3.**
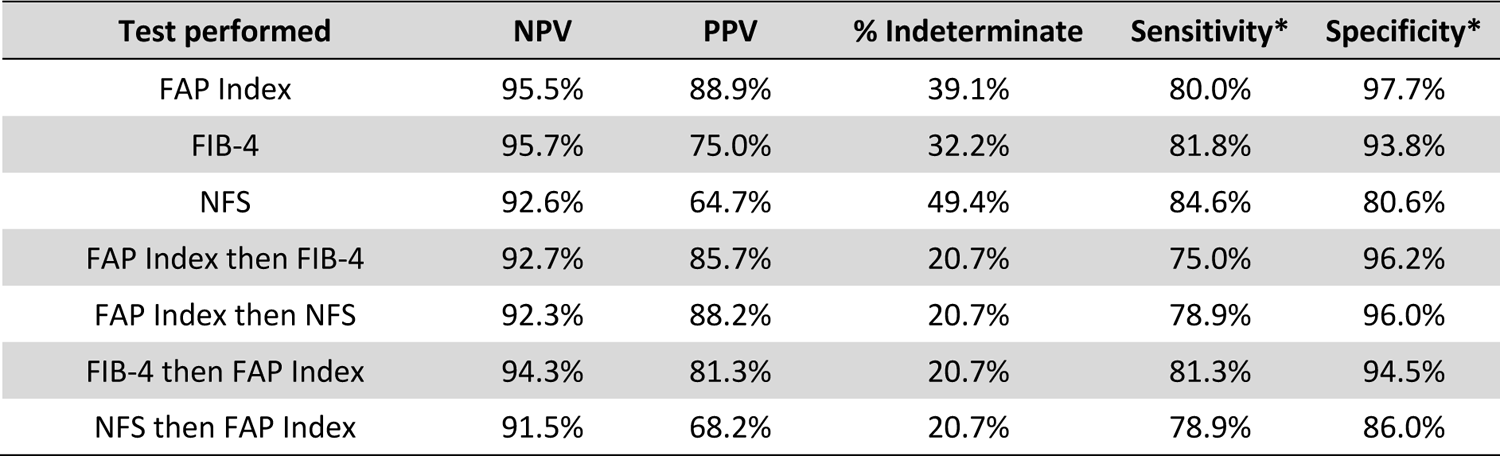
Summary table for these classification analyses of all tests in the training subcohort (n=87), reported for NPV, PPV, proportion of indeterminate, sensitivity and specificity. *: Sensitivity and Specificity were calculated with the indeterminate results excluded.

### 3.6 FAP Index validation

The AUROC for FAP Index to predict advanced fibrosis in the validation cohort was 0.841 (figure S5). In the validation cohort, FAP Index and FIB-4 produced similar outcomes, with specificity above 98%, which were superior to NFS (specificity 64.4%). The NPVs for all three tests were above 90% (figure 4A-C, table 4). The FIB-4 sensitivity was 37.5%, while sensitivities for FAP Index and NFS were over 50%. Importantly, all tests exhibited large proportions of indeterminate: 32.6% for FAP Index, 24.1% FIB-4 and 47.5% for NFS. FAP Index was significantly correlated with FIB4 and NFS in the validation cohort (P < 0.0001; supplemental figure 4).

**Figure 4.**
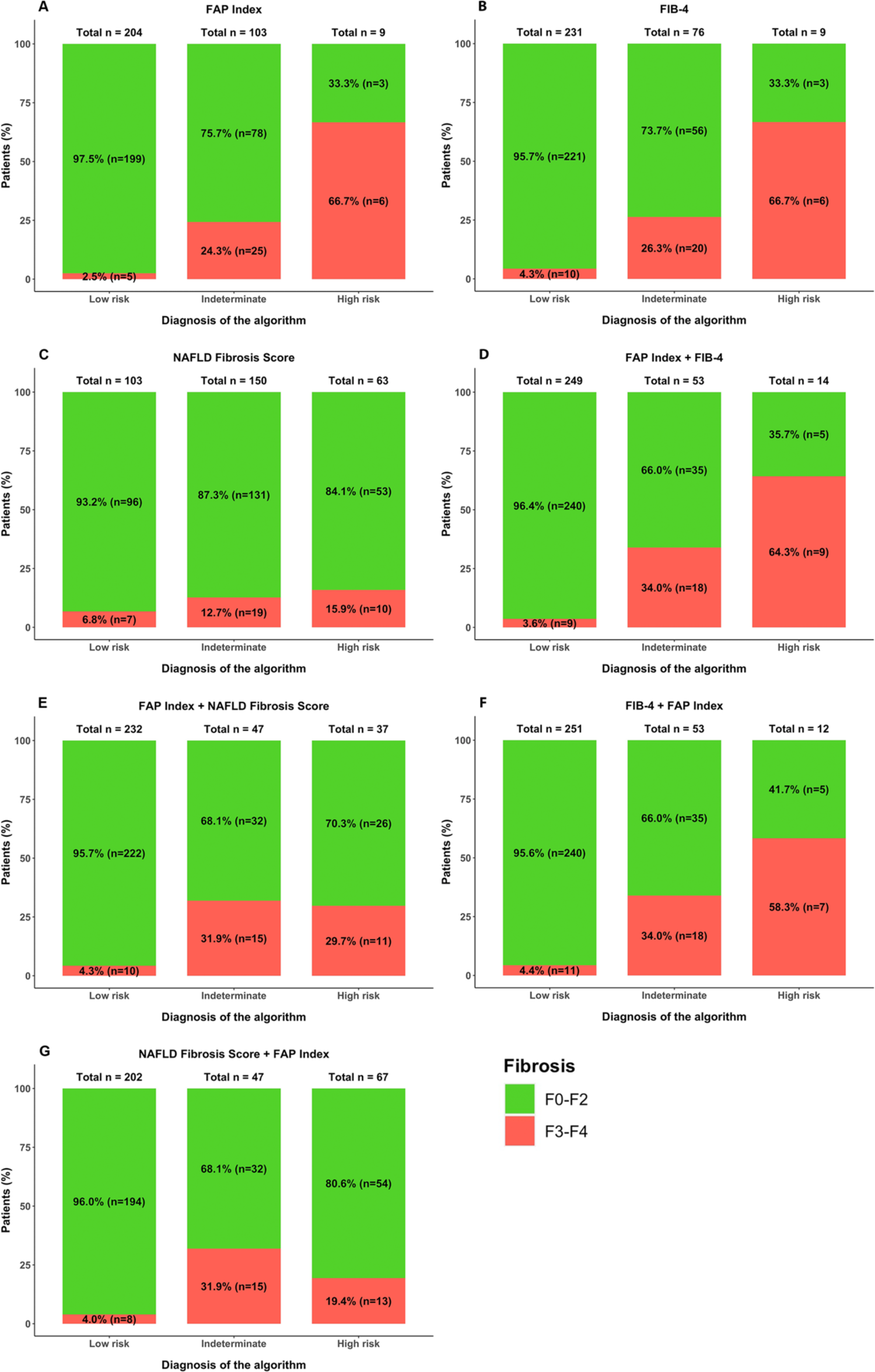
Classification analysis in the validation cohort. Fibrosis stages classification as a single test or sequential tests, as a function of subgroups defined by **A.** FAP Index classification. **B.** FIB-4. **C.** NAFLD Fibrosis Score. **D.** FAP Index followed by FIB-4. **E.** FAP Index followed by NAFLD Fibrosis Score. **F.** FIB-4 followed by FAP Index. **G.** NAFLD Fibrosis Score followed by FAP Index.

**Table 4.** Summary table for these classification analyses of all tests in the validation cohort, reported for NPV, PPV, proportion of indeterminate, sensitivity and specificity. * Sensitivity and Specificity were calculated with the indeterminate results excluded.

| Test performed | NPV | PPV | % Indeterminate | Sensitivity* | Specificity* |
| --- | --- | --- | --- | --- | --- |
| FAP Index | 97.5% | 66.7% | 32.6% | 54.5% | 98.5% |
| FIB-4 | 95.7% | 66.7% | 24.1% | 37.5% | 98.7% |
| NFS | 93.2% | 15.9% | 47.5% | 58.8% | 64.4% |
| FAP Index then FIB-4 | 96.4% | 64.3% | 16.8% | 50.0% | 98.0% |
| FAP Index then NFS | 95.7% | 29.7% | 14.9% | 52.4% | 89.5% |
| FIB-4 then FAP Index | 95.6% | 58.3% | 16.8% | 38.9% | 98.0% |
| NFS then FAP Index | 96.0% | 19.4% | 14.9% | 61.9% | 78.2% |

### 3.7 Sequential application of NITs

Next, sequential application of each possible combination of NITs was performed, with the primary aim to minimize the proportion of patients who were indeterminate without compromising diagnostic accuracy (figure 5). All combinations and sequences of NITs generated similar NPVs, above 90%, in both training cohort (table 3) and validation cohort (table 4). In the training cohort, FAP Index applied as either the 1^st^ line or 2^nd^ line NIT in combination with either NFS or FIB-4 reduced the proportion of indeterminate to 20.7% (figure 3D-G; table 3). In the validation cohort, FAP Index used in combination with NFS resulted in 14.9% indeterminates, whereas FAP Index with FIB-4 in any combination led to 16.8% indeterminates (table 4, figure 4D-G). In both cohorts, the relative reduction in the indeterminate population caused by adding the second line test reached 50%. However, combinations of FAP Index with FIB-4 followed by FAP Index maintained specificity of ∼95% in both cohorts (98% in validation cohort) compared to NFS combinations with FAP Index where specificity was as low as 78% in the validation cohort (tables 3 and 4). This is reflected in figure 4, where although NFS combinations classified more patients as high risk, up to 80% did not have histologic advanced fibrosis. Importantly, applying a step-wise test approach did not influence the sensitivity and PPV compared with applying a single test. Sensitivity was ∼75% in the training cohort, and ∼50% in the validation cohort. As expected in the validation cohort where the prevalence of advanced fibrosis was lower, NPV increased and PPV reduced compared to the training sub-cohort. Overall, the combination of FAP Index followed by FIB-4 generated the optimal diagnostic metrics, sufficiently reducing indeterminate results while maintaining an excellent NPV and acceptable PPV compared to single NITs.

**Figure 5.**
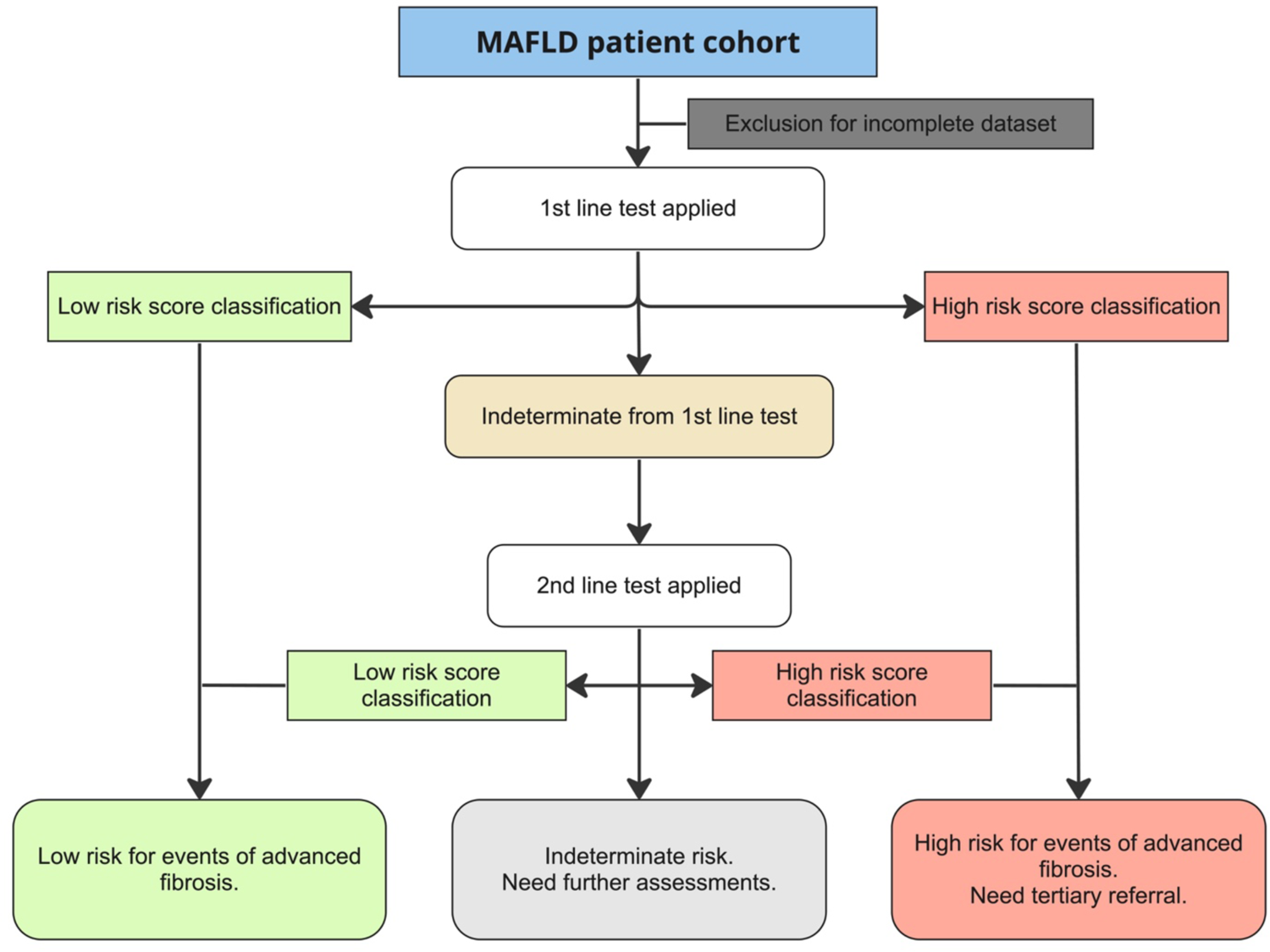
Flowchart for model validation in all cohorts, using two sequential tests to further classify the indeterminate patients from the 1^st^ line screening test.

## 4 Discussion

In this study, we demonstrated that circulating fibroblast activation protein (cFAP), a marker of activated stellate cells and myofibroblasts, can be incorporated into an algorithm and applied following NFS or FIB-4 to greatly diminish the number of indeterminate outcomes without compromising sensitivity or specificity for advanced hepatic fibrosis. This novel NIT, which we call FAP index, was examined in a training cohort and validated in a pooled cohort of individuals with metabolic fatty liver disease with variable prevalence of biopsy proven advanced fibrosis, type 2 diabetes, obesity and metabolic syndrome. The FAP Index utilises cFAP and three routinely obtained parameters; age, T2DM status and ALT. We have demonstrated the combination of FAP Index followed by FIB-4 provides a significant reduction in indeterminate results without compromising diagnostic accuracy.

Of the variables included in FAP Index, ALT is a well-established biomarker for the severity of liver injury; age is a known risk factor for progression for liver disease,^42^ T2DM and insulin resistance are drivers of metabolic fatty liver disease progression, and insulin resistance increases intrahepatic oxidative stress^3, 5, 43, 44^. The presence of these variables suggests a dysmetabolic state with liver injury resulting in continual hepatic stellate cell (HSC) activation. We have shown that FAP is strongly associated with the presence of advanced fibrosis^20–22, 25^, and FAP is found in and on activated HSC and myofibroblasts and contributes to collagen turnover^20, 21^. Moreover, new FAP/^42^radionuclide-based 3D imaging methods have shown that intrahepatic FAP strongly aligns with fibrosis severity^10^. Serum FAP drops following liver transplantation^26^, which suggests that liver is the origin of increased circulating FAP in patients with advanced fibrosis.

In the training sub-cohort, FAP Index showed excellent accuracy in predicting the presence of advanced fibrosis, with an AUROC of 0.875 (95% CI: 0.813-0.938), while FIB-4 and NFS achieved comparable AUROCs of 0.899 (95% CI: 0.833-0.965) and 0.781 (95% CI: 0.672-0.89) respectively. Our findings demonstrated that the FAP Index has a strong discriminating capability among cohorts of varied patient composition and prevalence of advanced fibrosis. However, the percentage classified as indeterminate was relatively large as standalone NIT, which is an established issue for FIB-4 and NFS.

In contrast, sequentially using FAP Index with either FIB-4 or NFS was shown to dramatically reduce the number of indeterminate results by between 30% and 70%, depending on the NIT combination. ^13, 45^ Notably, the two-step strategy did not significantly alter the NPV, PPV nor specificity when using FAP Index as either 1^st^ line test or 2^nd^ line test in combination with either FIB-4 or NFS. Indeed, our group and others have shown that sequential screening tests that employ dual cut-offs can the increase diagnostic accuracy of NITs and thereby reduce the number of biopsy referrals^46–48^. Similarly, a recent evaluation of the current EASL algorithm by Patel and colleagues found that a stepwise strategy can improve diagnostic accuracy^8^. However, the use of FIB-4 or NFS as first line NITs resulted in approximately 35% and up to 65% indeterminate results, respectively, in both the diabetes clinic and primary care cohorts analysed. Furthermore, up to 84% needed further assessment (ie indeterminate or high-risk classification) when using NFS or FIB-4 ^8^. In contrast, our two-step algorithm of FAP Index followed by FIB-4 resulted in 39% in the training subcohort and only 21% in the validation cohort who would need further investigation by the same criteria (ie indeterminate or high-risk classification). This reduction compared to the Patel study was in the context of similar advanced fibrosis prevalence in our training subcohort of 27%, however only 48 patients underwent biopsy in the Patel cohorts.

Another advantage of the sequential combination of FAP Index with FIB-4 was the high specificity. It is essential to make sure that false positives and, particularly in the context of screening, that false negatives are minimized. Our validation cohort had a prevalence of advanced fibrosis similar to the community of 11%^35, 36^ and using FAP Index followed by FIB-4 resulted in 3.6% (9/249) false negatives and 35.7% (5/14) false positives. This strategy provided the lowest rate of false negative diagnoses and comparable false positives to FIB-4 and NFS, in addition to the dramatic reduction in indeterminate results. This suggests that FAP Index has potential benefit in community screening algorithms over current standard of care.

FAP Index has potential as a readily applicable first line test as it contains variables that are easily obtained, and cFAP measurement requires only a standard fluorescence measurement device, so its cost is likely to be comparable with AST and ALT. Similarly, the measurement of ordinal cFAP in FAP Index could be readily modified into a point-of-care lateral flow test, or as a reflex test during the measurement of liver biochemistry as part of clinical pathology services.

Community demand for screening for hepatic fibrosis is rising, particularly in diabetes clinics, so inexpensive, simple liver fibrosis testing is needed. Although biopsy is considered as the most accurate diagnostic tool in clinical guidelines, it has unavoidable pitfalls such as accessibility, invasiveness, the need for specialised staff, variability between practitioners, and economic impacts. While elastography is a useful alternative to biopsy, accuracy drops when BMI exceeds 44, and it is influenced by operator-dependent variables and barriers still exist in terms of accessibility and/or affordability^49^. Therefore, the potential for FAP Index as an accurate blood test and point-of-care test in routine screening could alleviate total health care costs. More importantly, the enzyme activity of soluble FAP was found to have high stability with extended ultracold storage ^24^ (supplemental figure2), which means that the cFAP assay is amenable as a subsequent test after an initial FIB4 or NFS is completed. Thus, FAP Index as a reflex test would not require an additional blood collection, which further increases convenience and reduces health care expenditures.

Following a successful clinical trial^50^, the U.S. Food and Drug Administration recently approved a thyroid hormone receptor beta (THR-β) agonist^51^ as the first therapy for non-cirrhotic steatohepatitis and advanced stages of liver fibrosis. This further motivates the need for more accurate triage of patients with advanced fibrosis. NFS and FIB-4 are currently regarded as standard of care for fibrosis screening, however, these NITs rely upon parameters that are strongly associated with liver function, liver injury and inflammation, rather than directly with fibrosis per se^11, 12^. As a result, these tests can yield large proportions of misclassifications among patients at high risk of metabolic fatty liver disease^14^. Liver fibrosis is dependent on the intensity of tissue remodelling and myofibroblast activation. Therefore, including a simple biomarker such as FAP that directly reflects the fibrotic process is advantageous.

A major strength of this study was the robust training and validation cohorts. The prevalence of fibrosis in the training cohort was similar to that of hepatology clinic populations, while the validation cohort mimics community screening in an at-risk population, albeit with high frequencies of obesity and diabetes. We have demonstrated that our FAP assay is robust and activity can be measured via a robust and simple one-step enzyme assay. We used validated methodology to arrive at the FAP Index and have performed comprehensive statistical analysis to examine the influence of using different NITs alone or in combination. The study limitations were sample selection, sample size and non-centralised histologic fibrosis assessment. Comparison of NITs in the training cohort were only available in a subset of patients, however, the findings were replicated in the validation cohort.

## 5 Conclusion

We have developed and validated FAP Index; a simple and novel non-invasive test for triaging the risk of advanced fibrosis in metabolic associated fatty liver disease. FAP Index demonstrated excellent diagnostic accuracy with an AUROC of 0.875 as a standalone test. However, sequential use of FAP Index followed by FIB-4 yielded more favourable results with a reduction of indeterminately classified patients by one third compared to FIB-4 alone in both training and validation cohorts. This was achieved while maintaining an NPV of >95% and having the lowest percentage of false negative diagnoses of any NIT strategy examined. Moreover, FAP Index used sequentially with NFS compared to NFS alone generated even more striking results. FAP Index includes a direct marker of fibrogenesis (cFAP), which is simple and inexpensive to measure with potential application as a point of care or reflex test. Therefore, FAP Index demonstrates several advantages over the current first-line standard of care NITs for triaging risk of advanced fibrosis due to MAFLD and should be developed for clinical use.

## ABBREVIATIONS

ALP: alkaline phosphatase

ALT: Alanine transaminase

APRI: AST to Platelet Ratio Index; AST to Platelet Ratio Index

AST: Aspartate transaminase

AUROC: area under the receiver operating characteristics curve

BMI: body mass index

cFAP: circulating fibroblast activation protein

ECM: extracellular matrix

ESAL: equivalent single axle load

FAP: fibroblast activation protein

FGF-21: fibroblast growth factor 21

GGT: Gamma-glutamyl transferase

HSC: hepatic stellate cell

IQR: inter-quartile range

MAFLD: metabolic associated fatty liver disease

MRE: magnetic resonance elastography

NFS: NAFLD Fibrosis Score

NIT: non-invasive test

NPV: negative predictive value

PLT: platelet

PPV: positive predictive value

ROC: receiver operating characteristics curve

T2DM: type 2 diabetes mellitus.

## AUTHOR CONTRIBUTIONS

ZVW: study concept and design, statistical analysis, analysis and interpretation of data, drafting the manuscript; BBB: acquisition of data, review and manuscript editing, study supervision; TP: study concept, statistical analysis; WWB, SEP, JHSL, PVDV, JG, LAA: Supply study materials, critical revision of the manuscript; GO, ME: acquisition of study samples; KB: statistical analysis and interpretation of data; HEZ: study concept and design, statistical analysis; GWM: acquisition of study samples, obtained funding; AM: study concept and design, critical revision of the manuscript for important intellectual content, study supervision; MDG: study concept and design, obtained funding, critical revision of the manuscript for important intellectual content, study supervision.

## Patient and public involvement

Patients and/or the public were not involved in the design, or conduct, or reporting, or dissemination plans of this research.

**Patient consent** for this publication, Consent was not required from patients.

## Ethics approval

The approvals were HREC_X18-0241 in Sydney Local Health District, 2019/ETH02319 in Westmead Hospital, RGS01287 in Sir Charles Gairdner Hospital, and 195/15 in The Alfred Hospital. All participants provided written informed consent and their serum samples and information were handled in accordance with ethics approvals and attendant regulations of each hospital.

## Supplemental material

This content has been supplied by the authors.

## Data Availability

All data produced in the present study are available upon reasonable request to the authors

