## Supplemental material for "Development and validation of a novel circulating fibroblast activation protein - based predictive model to improve fibrosis risk stratification in metabolic liver disease population"

### Supplemental Methods

#### S1.1 | The FAP specificity of 3144-AMC

The FAP-specific substrate, 3144-AMC [1], was compared to substrates compound 4 (a structurally identical compound of 3144-AMC) [2] and compound 6c [2]. 3144-AMC, compound 4 and compound 6c were solubilised in DMSO then diluted to a stock concentration of 10 mM, aliquoted and stored at -20 °C until required. Prior to running the cFAP assay, a 1:20 dilution of the substrates was made in TE buffer with 30 µL pipetted into each well to a final concentration of 150 µM and final volume of 100 µL. To determine if the substrates hydrolysed other members of the DPP4 family, inhouse, purified recombinant DPP4 and FAP [3, 4] and recombinant prolyl oligopeptidase (PREP; R&D Systems Catalog number 4308-SE-010) were prepared in serial dilutions (0-10 ng) and run together with mouse and human samples.

### Supplemental Results

#### S2.1 | Enzyme assay specificity for FAP

We previously demonstrated the specificity of 3144-AMC (previously called ARI-3144-AMC) as a substrate for tissue derived mouse FAP and for circulating fibroblast activation protein alpha (cFAP) in human and mouse plasma and serum [1]. Furthermore, we discovered a ~20x increase in cFAP activity in cirrhotic liver compared to healthy liver and up to 20-fold more cFAP in mouse than in healthy humans. Here, we have optimised the FAP enzyme activity assay and further showed the specificity of 3144-AMC for FAP and that of compound 6c, which is also a specific substrate for FAP [2] (Fig. S2). 3144-AMC, compound 4 [2] and compound 6c exhibited similar hydrolysis by FAP (Fig. S2A, D). Compound 4 was hydrolysed by recombinant PREP, whereas no hydrolysis was observed with either 3144-AMC or compound 6c (Fig. S2B, E). Purified DPP4 did not detectably hydrolyse any of the three substrates (Fig. S2C). In the presence of human serum, all substrates exhibited similar FAP activity (Fig. S2F, G). Finally, we assessed substrate hydrolysis by natural FAP that is in the serum of wild-type and FAP enzyme negative mice (Fig. S2H-J). FAP activity measurements in the serum of wild-type mice were similar across all three substrates (Fig. S2H-I). Most convincing of the specificity of 3144-AMC for FAP was the lack of FAP activity detected in the serum of FAP enzyme negative mice (Fig. S2J), concurring with our previously published data [1]. Taken together, these data clarify that compound 6c and 3144-AMC have equal potency and specificity for measuring FAP activity. The chemical structures of compound 6c and 3144-AMC are identical except for two fluorine substituents on the proline ring in compound 6c. Therefore, the above data shows that this structural difference is inconsequential for their interaction with FAP. Compound 4 was synthesized to lack those fluorines and thus be identical to 3144-AMC [2]. Why compound 4 was not as specific as 3144-AMC is unclear but may have been caused by a degree of stereochemical impurity at the Ala position.

#### S2.2 | Enzyme stability and FAP assay reproducibility

In this study, FAP stability and the reproducibility of the cFAP enzyme assay on human serum samples was examined by repeat assay of the entire Westmead Hospital cohort five years after cFAP was first measured for the initial data collection. No statistical difference was observed between these two replicate assays (P = 0.11) (Fig. S3). By transforming the cFAP activity into ordinal levels using the cut-offs mentioned above, the histograms for these two replicate assays were almost identical (Fig. S3A). Moreover, when each cFAP ordinal dataset from the assays performed five years apart by different personnel using different batches of reagents were incorporated into the FAP Index calculation, no significant difference was identified (P = 0.70). This was consistent with our previous finding that freeze/thaw cycles do not affect cFAP activity [1], and strongly indicates the stability of cFAP activity in storage and a robust and very reproducible assay.

#### S2.3 | Correlation of FAP Index with established NITs

FAP Index was also examined for its correlation with currently established NITs. As shown in Supplementary table 2, FAP Index was significantly correlated with both FIB-

4 and NFS ($P < 0.001$). In addition, FAP Index exhibited a linear and positive regression with both FIB-4 and NFS across all study cohorts (Fig. S4). In the training cohort, the goodness-of-test analysis showed that FAP Index moderately correlated with both FIB-4 Index ($r2 = 0.25$) and NFS ($r2 = 0.23$). In the validation cohort, FAP Index correlated with FIB-4 Index ($r2 = 0.41$, $r2 = 0.22$, respectively), and with NFS ($r2 = 0.2$, $r2 = 0.04$, respectively).

### Supplemental Tables

***Table S1. Patient characteristics for each individual cohort***. *Statistically significant differences among all three cohorts (P-value). Statistically significant differences between training cohort and Alfred Hospital cohort or Westmead Hospital cohort were obtained by post hoc (Tukey's) multiple comparison tests and are indicated by asterisks. Data presented as median ± IQR for continuous variables.*

| | ***Training cohort (n=160)*** | ***Validation cohort (n =332)*** | | |
|---|---|---|---|---|
| | ***Training cohort (n=160)*** | ***Alfred cohort (n=182)*** | ***Westmead cohort (n = 150)*** | ***P-value*** |
| Age [a] | 52±18.25 | 45±19.75**** | 52±16 | <0.0001 |
| Gender (Male) [b] | 58 (36%) | 44 (24%) | 76 (50.7%) | <0.0001 |
| Diabetes (1) [b#] | 57 (36%) | 41 (23%) | 57 (38%) | 0.004 |
| BMI [a] | 38.19±12.05 | 45.15±10.86**** | 30.82±7.48**** | <0.0001 |
| ALT (U/L) [a] | 40±42.25 | 33±26* | 66.5±51** | <0.0001 |
| AST (U/L) [a] | 32.5±20 | 27±13* | 55.58±29**** | <0.0001 |
| GGT (U/L) [a] | 40±60.25 | 33±21**** | 85.5±94.5 | <0.0001 |
| PLT (x$10^9$/L) [a] | 225±94 | 238.5±88.25 | 241±88.7 | 0.13 |
| ALP (U/L) [a] | 81±35.5 | 69±27 | Nd | Nd |
| Insulin (mU/L) [a] | 11±15 | 7.1±7.8** | 15±12 | 0.002 |
| Fibrosis staging [b] | | | | <0.0001 |
| F0 | 81 | 139 | 41 | |
| F1 | 35 | 36 | 48 | |
| F2 | 10 | 3 | 27 | |
| F3 | 17 | 3 | 26 | |
| F4 | 12 | 1 | 8 | |
| Advanced fibrosis [b] | 32 (20%) | 4 (2%) | 34 (22.7%) | <0.0001 |
| cFAP activity (pmol AMC/min/L) [a] | 995.74±579.3 | 1193.22±434.04** | 1419±635.1**** | <0.0001 |
| Ordinal cFAP [b] | | | | <0.0001 |
| Level 0 | 38 | 11 | 4 | |
| Level 1 | 94 | 142 | 87 | |
| Level 2 | 28 | 29 | 59 | |

Notes: * p-value<0.05, ** p-value<0.01, *** p-value<0.001, **** p-value<0.0001 of Mann-Whitney t-test comparison with cohort P.

[a] One-way ANOVA was used to identify statistically significant difference among means across all cohorts.

[b] Chi-square test was used to identify statistically significant difference among proportion across all cohorts.

#: The value 1 was assigned to individuals with type 2 diabetes.

***Table S2.*** *Correlation analyses between circulating FAP activity and each other parameter in each cohort. Data presented as Pearson coefficient (P-value). #: The value 1 was assigned to individuals with type 2 diabetes.*

| | ***Training cohort*** | ***Validation cohort*** |
|---|---|---|
| Age | 0.13 (0.11) | 0.12 (0.03) |
| Gender (Male) | 2050 (0.001) | 9337 (0.01) |
| T2DM # (1) | 3396 (0.10) | 11157 (0.74) |
| Hypertension | 3608 (0.14) | 13104 (0.56) |
| Weight (kg) | -0.04 (0.59) | 0.02 (0.82) |
| Height (m) | 0.10 (0.22) | 0.19 (0.009) |
| BMI | -0.11 (0.15) | -0.25 (<0.001) |
| Bilirubin (umol/L) | 0.11 (0.15) | 0.13 (0.08) |
| ALP (U/L) | 0.24 (0.002) | 0.13 (0.08) |
| ALT (U/L) | 0.28 (<0.001) | 0.19 (<0.001) |
| AST (U/L) | 0.36 (<0.001) | 0.18 (0.001) |
| GGT (U/L) | 0.34 (<0.001) | 0.13 (0.02) |
| AST/ALT | 0.02 (0.83) | 0.02 (0.7) |
| Alb (g/dL) | -0.14 (0.08) | 0.17 (0.001) |
| PLT (x$10^9$/L) | -0.28 (0.004) | -0.06 (0.29) |
| Creatinine (umol/L) | 0.10 (0.35) | - |
| Glucose (mmol/L) | 0.004 (0.95) | -0.09 (0.26) |
| Insulin (mU/L) | 0.31 (<0.001) | 0.18 (0.001) |
| HOMA-IR | 0.28 (0.002) | 0.16 (0.004) |
| TG (mmol/L) | 0.04 (0.66) | 0.16 (0.003) |
| Steatosis | 5.96 (0.11) | 7.86 (0.049) |
| Inflammation | 3.45 (0.18) | 7.54 (0.05) |
| Ballooning | 9.10 (0.01) | 9.12 (0.01) |
| Fibrosis | 26.95 (<0.001) | 37.52 (<0.001) |

***Table S3***. *Baseline characteristics of the training cohort and the training subcohort. Statistical significance was examined by Mann-Whitney U test.*

| | ***Training cohort (n=160)*** | ***Training subcohort cohort (n=87)*** | ***P-value*** |
|---|---|---|---|
| Age | 52±18.25 | 57±15 | 0.03 |
| Gender (Male) | 58 (36%) | 34 (39.1%) | 0.76 |
| Diabetes (1) # | 57 (36%) | 34 (39.1%) | 0.69 |
| BMI | 38.19±12.05 | 34±11.4 | 0.02 |
| ALT (U/L) | 40±42.25 | 51±45.5 | 0.22 |
| AST (U/L) | 32.5±20 | 37±26.5 | 0.15 |
| GGT (U/L) | 40±60.25 | 61±97.5 | 0.02 |
| Platelets (PLT; x$10^9$/L) | 225±94 | 216±101.5 | 0.57 |
| ALP (U/L) | 81±35.5 | 92±51 | 0.04 |
| Insulin (mU/L) | 11±15 | 15±17 | 0.02 |
| HOMA-IR | 0.4 (0.76) | 0.68 (0.85) | 0.01 |
| Fibrosis staging | | | 0.68 |
| F0 | 81 (51.3%) | 38 (43.7%) | |
| F1 | 35 (22.2%) | 19 (21.8%) | |
| F2 | 10 (6.3%) | 6 (6.9%) | |
| F3 | 17 (10.8%) | 11 (12.6%) | |
| F4 | 15 (9.5%) | 13 (14.9%) | |
| Advanced fibrosis | 32 (20.3%) | 24 (27.6%) | 0.25 |
| cFAP activity (pmol AMC/min/L) | 995.74±579.3 | 1075±616.5 | 0.56 |
| cFAP activity ordinal | | | 0.54 |
| Level 0 | 38 (23.8%) | 21 (24.1%) | |
| Level 1 | 94 (58.5%) | 46 (52.9%) | |
| Level 2 | 28 (17.5%) | 20 (23%) | |

Notes: p-value < 0.05 indicates statistical significance. Data presented as median ± IQR for continuous variables, and prevalence (%) for categorical variables. #: The value 1 was assigned to individuals with type 2 diabetes.

### Supplemental Figures

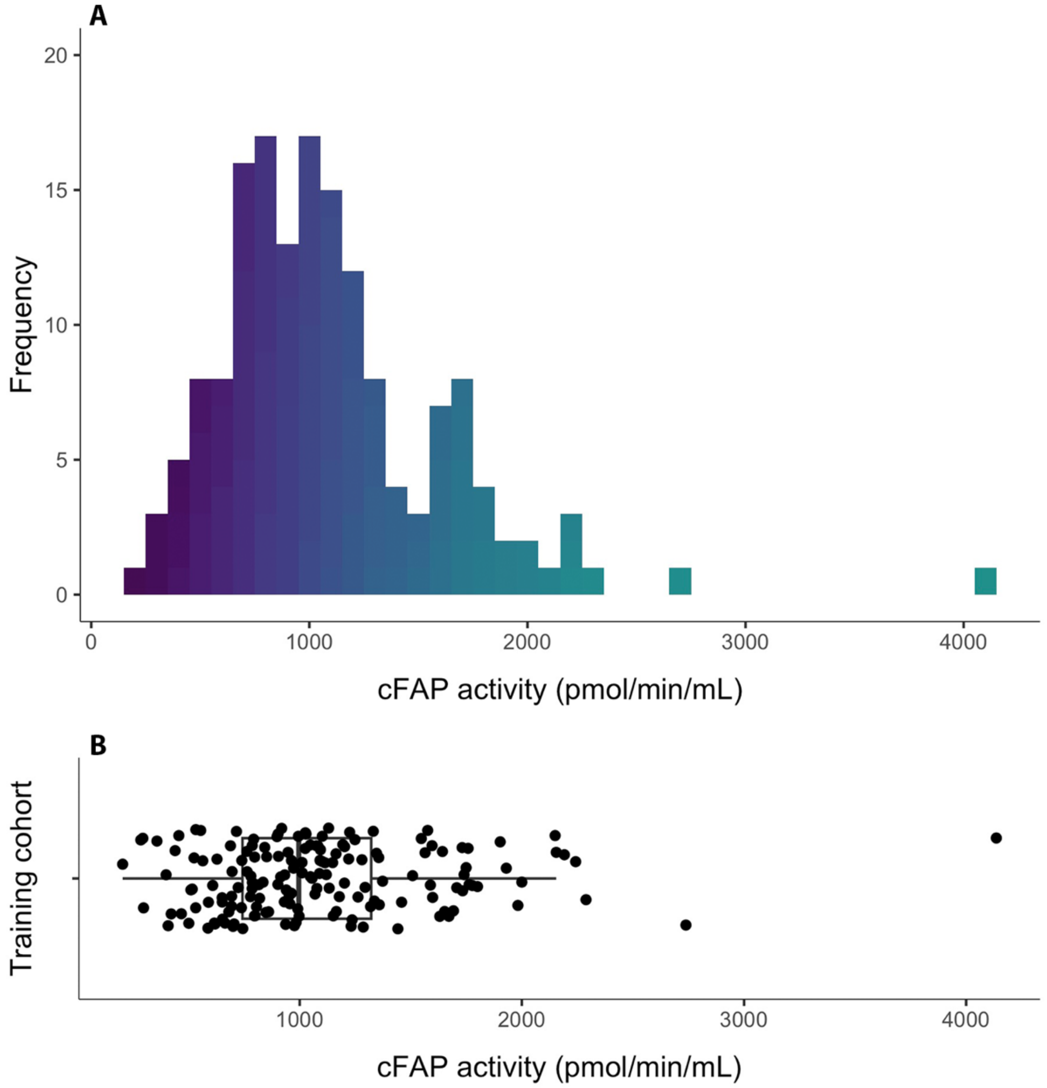

***Figure S1. A).*** *Histogram of cFAP activity in the training cohort.* ***B)****. Boxplot of cFAP activity in the training cohort .*

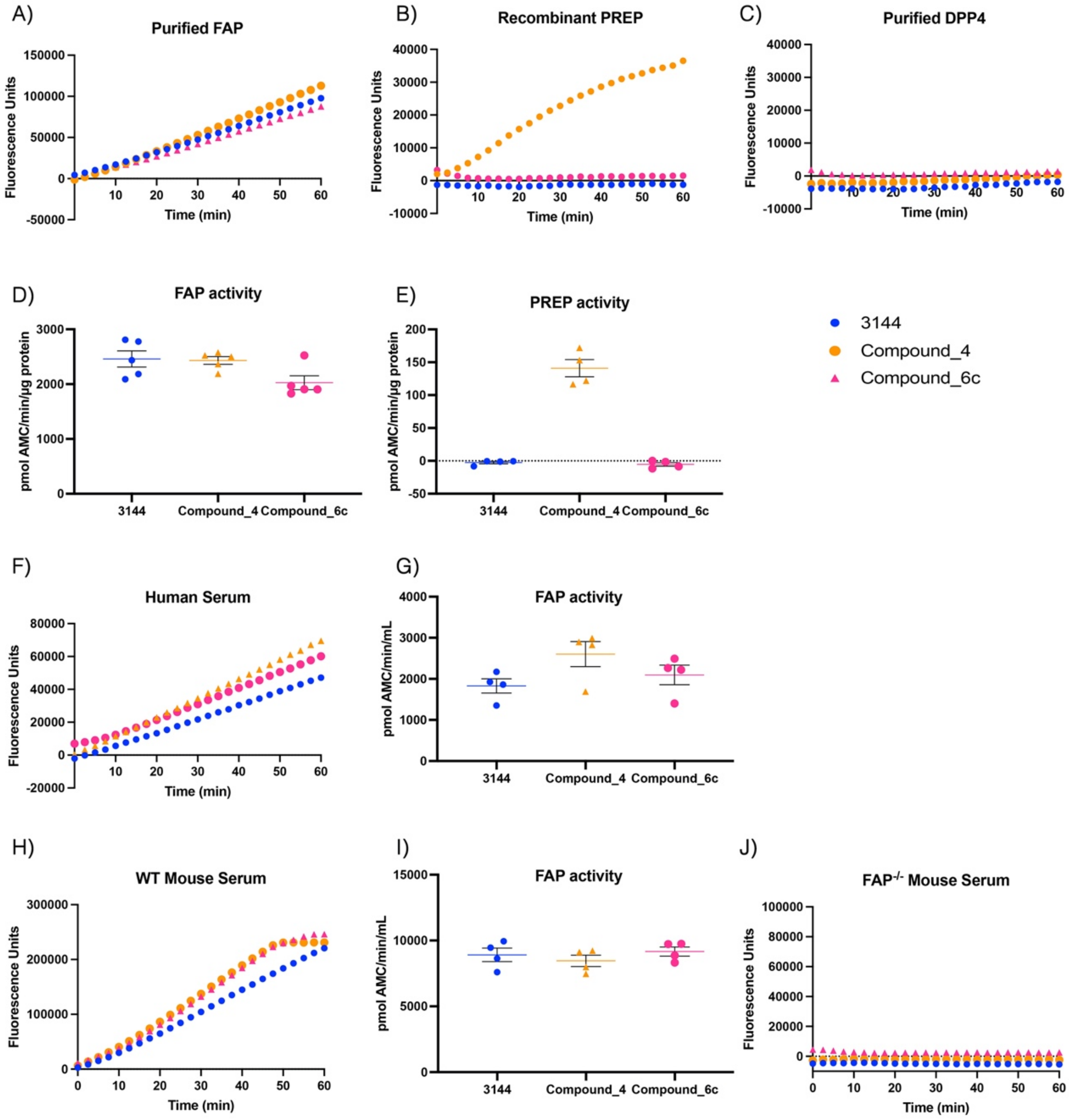

***Figure S2. 3144-AMC is a specific substrate of fibroblast activation protein alpha (FAP).***

An in-house enzyme-based assay was used to examine the hydrolysis (A) purified FAP, (B) recombinant PREP and (C) purified DPP4 in the presence of 3144-AMC (blue), compound 4 (orange) and compound 6c (pink). Activity of (D) FAP (n=5) and (E) PREP (n=5) was calculated on the linear segment of fluorescence. All three substrates were then used to determine hydrolysis of human serum (F) and subsequent FAP activity (G) (n=5). The three substrates were also used to measure hydrolysis and FAP activity in the serum of wild-type mice (H, I) (n=4) and FAP enzyme negative mice (J) (n=4). Fluorescence was measured every 2.5 minutes for 1 hour at

37 °C in a plate reader with excitation at 355 nm and emission at 450 nm. Data presented as mean ± SEM.

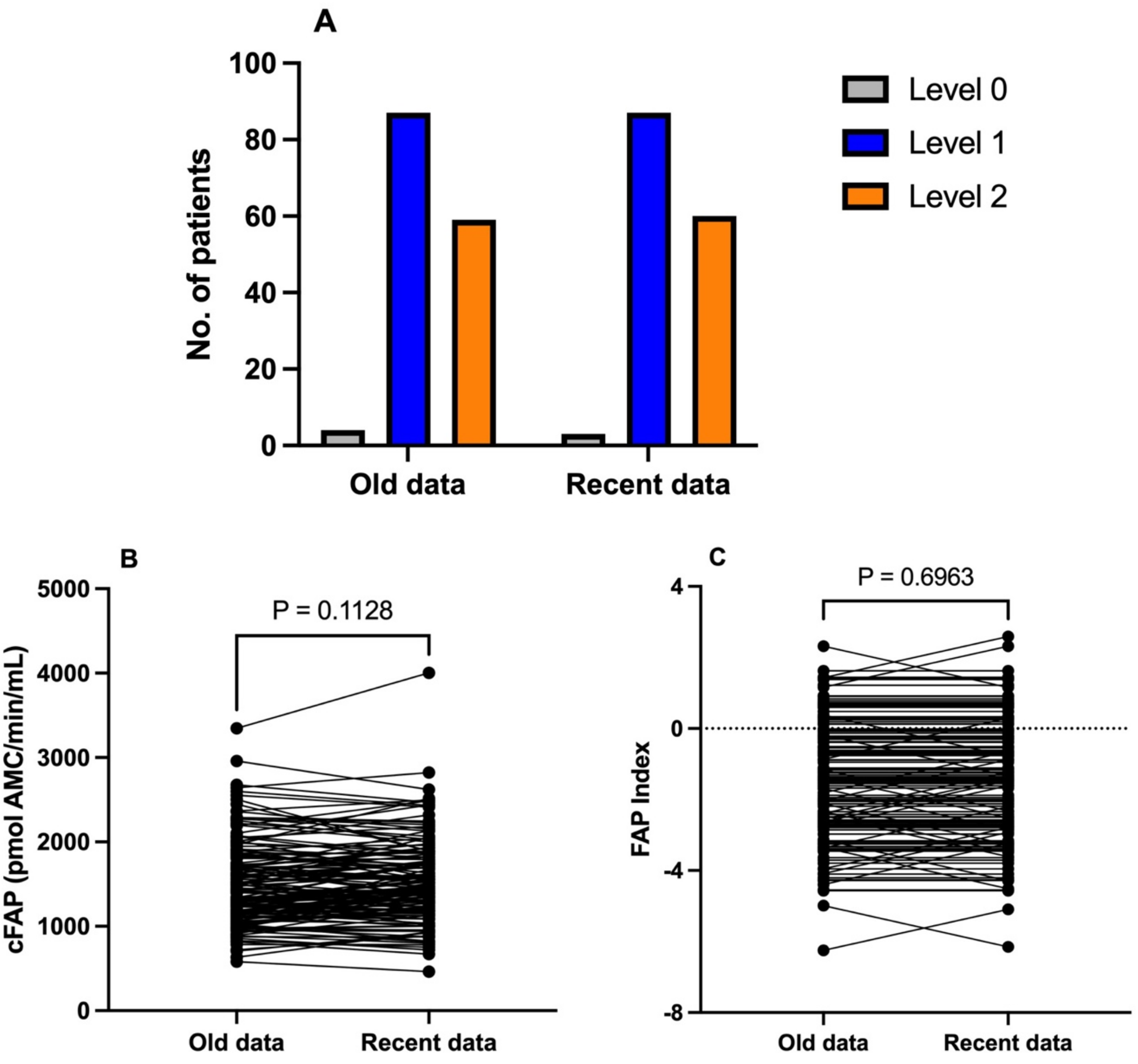

***Figure S3***. *Pairwise analysis of cFAP activity measured at two different time points and performed by two different persons on serum samples from 150 patients of Westmead Hospital. A) The histogram of cFAP activity transformed into the ordinal levels 0, 1 and 2. The independent chi-square test showed no significant difference in the distribution of the ordinal rank ($X^2$ = 0.1513, P-value = 0.93). B) Paired t-test showed no significant difference between the two measurements ($t_{149}$ = 1.595, P-value = 0.11). C) Paired t-test showed no significant statistical difference between the two measurements of FAP Index calculated using the old and new cFAP activity measurements ($t_{149}$ = 0.39, P-value = 0.70).*

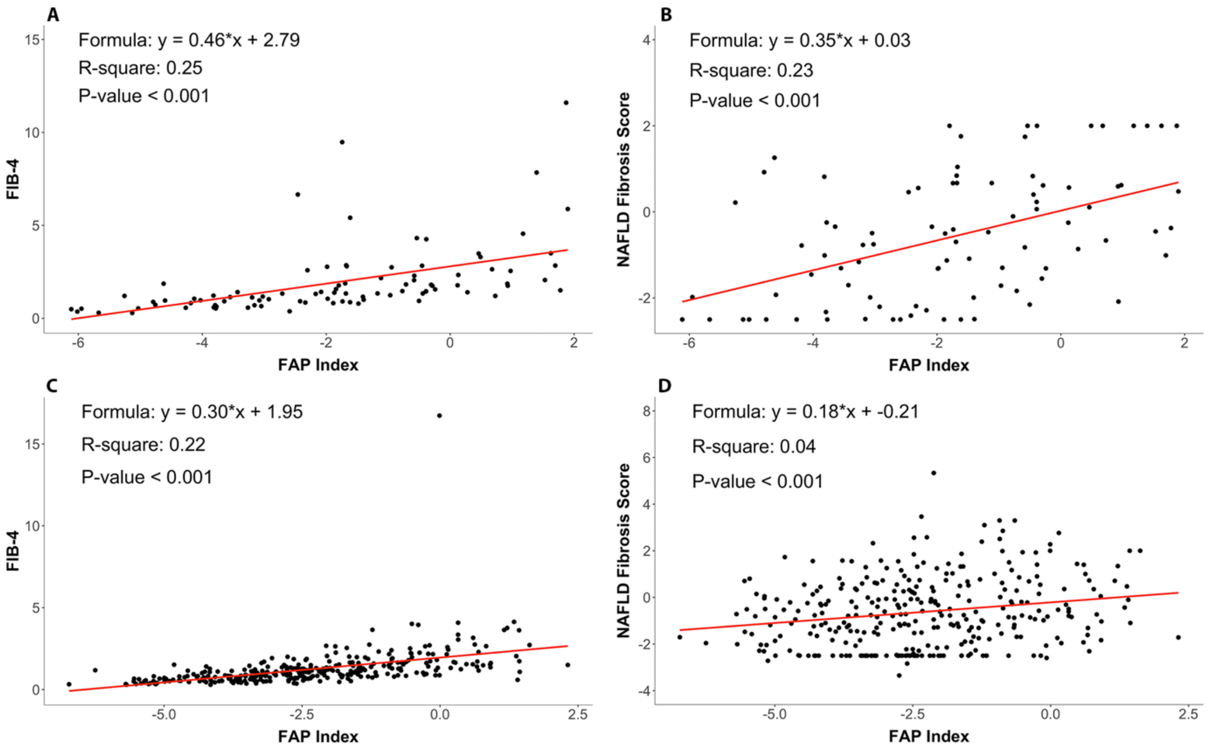

***Figure S4.*** *Correlation analyses of FAP Index with FIB-4 (left) and with NAFLD Fibrosis Score (right) in the training cohort* ***(A, B)*** *and the validation cohort* ***(C, D)****.*

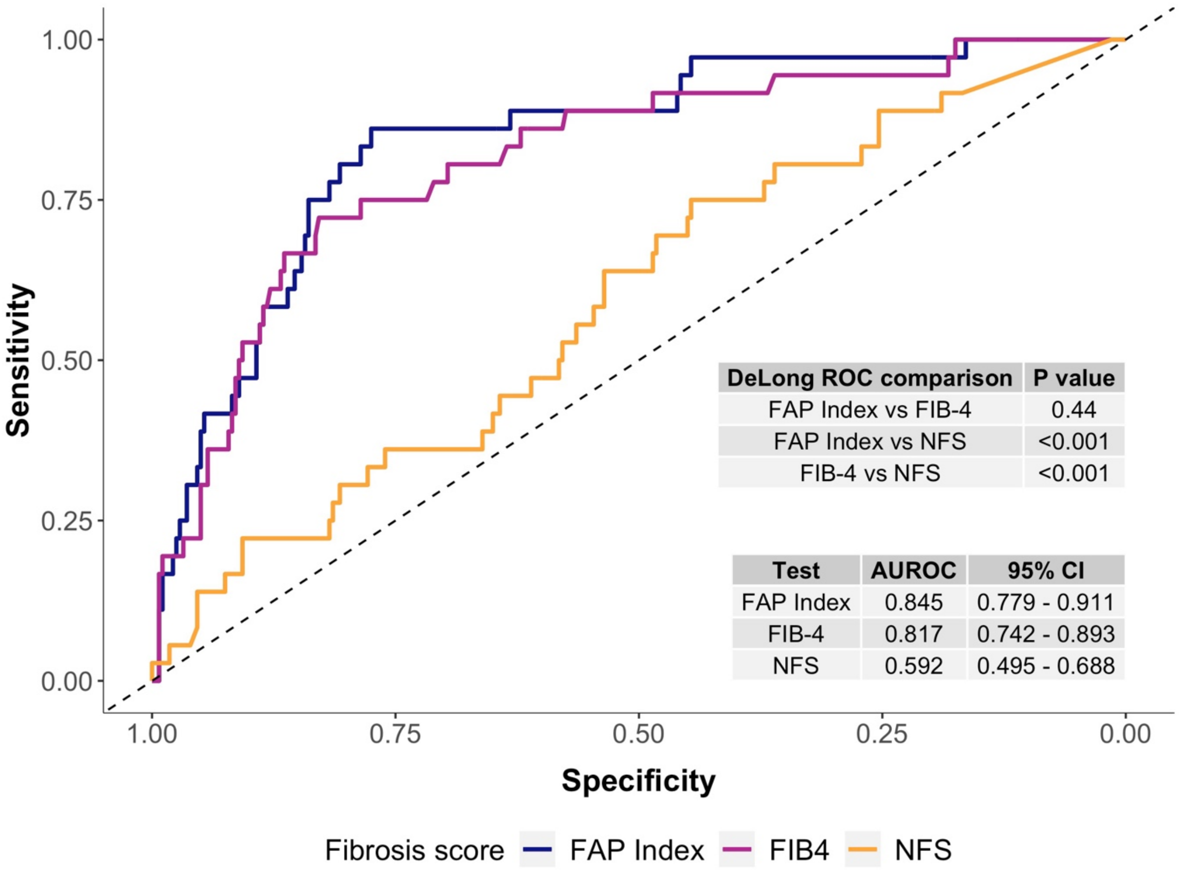

***Figure S5.*** *The ROC curves for established NITs to illustrate the overall discriminative performance in the validation cohort. CI: confidence interval.*
